# High prevalence of mixed *Plasmodium falciparum* and *Plasmodium vivax* co-infections in Duffy negative individuals in Ethiopia

**DOI:** 10.1101/2025.09.16.25335845

**Authors:** Baura Tat, Lionel Brice Feufack-Donfack, Bernis Neneyoh Yengo, Virak Eng, Tassew Tefera Shenkutie, Abnet Abebe, Meshesha Tsigie, Costanza Tacoli, Nimol Khim, Sindew M. Feleke, Eugenia Lo, Jean Popovici

## Abstract

Malaria remains a major public health concern in Ethiopia, where both *Plasmodium falciparum* and *Plasmodium vivax* are endemic. Notably, *P*.*vivax* infections have been documented in duffy-negative individuals, who were traditionally thought to be resistant to this parasite. *PvDBP* gene duplication has been proposed as a mechanism for P. *vivax* to invade erythrocytes in these individuals, but evidence is limited.

We conducted a cross-sectional study from 2020 to 2021 with1,154 febrile patients in three Ethiopian regions: Gambela, Oromia, and SNNPR. *Plasmodium* species were identified by PCR, Duffy genotypes by TaqMan allelic discrimination assay and *PvDBP* gene duplication by semi-nested PCR.

*Plasmodium* infection was detected in 46% of patients: 49% had *P. falciparum*, 26% had mixed *P*.*falciparum*/ *P*.*vivax* infections, and 23% had *P. vivax* mono-infections. Duffy genotyping showed 41% were Duffy negative, 44% heterozygous, and 16% Duffy positive homozygous, with significant regional differences. As expected, *P. vivax* mono-infections were rare among Duffy-negative individuals (2.4%), while *P. falciparum* infection prevalence was similar across Duffy genotypes. Among *P. falciparum*/*P. vivax* mixed-infections, 25% were Duffy negative. Off 155 samples successfully tested for the *PvDBP* gene duplication, 78% showed gene duplication, especially in Gambela (94%), where Duffy-negative individuals were most prominent. The prevalence of duplication did not differ between Duffy-positive homozygous and heterozygous individuals. Only one *P. vivax* mono-infection in a Duffy negative individual was examined and no gene duplication was detected.

Our findings suggest possible interactions between Duffy negative infection by *P. vivax* and co-infection with *P. falciparum*. The high prevalence of *PvDBP* gene duplication, particularly in regions with high Duffy negativity, may reflect evolutionary pressure to overcome host receptor barriers. These results underscore the need for molecular surveillance to monitor emerging parasite adaptations in populations previously considered resistant to *P. vivax*.

## Introduction

Malaria is one of the most prevalent and potentially lethal infectious diseases globally, accounting for about 249 million cases and 608,000 fatalities in 2022. *Plasmodium vivax* (*P. vivax*) and *Plasmodium falciparum (P. falciparum)* account for most of the malaria cases (6.9 million and 242.1 million cases, respectively) worldwide [1]. Even though there has been progress towards malaria elimination in the past years, the disease remains a major cause of mortality and morbidity in African countries. In Ethiopia, *P. vivax* and *P. falciparum* account for roughly 33% and 67% of all malaria cases, respectively, and it is one of the few African countries where *P. vivax* remains persistently prevalent [2].

It is well established that the Duffy Antigen Receptor for Chemokines (*DARC*) is the main receptor for *P. vivax* parasites, enabling erythrocyte invasion following interaction with the *P. vivax* Duffy Binding Protein (*PvDBP*) [3,4,5]. *DARC* is expressed on all erythrocytes, including reticulocytes, the target host cells of *P. vivax*. A single point mutation located in the GATA-1 transcription factor binding site of the *DARC* gene promoter (− 67 T>C) suppresses the expression of this receptor in erythroid cell lines, resulting in a Duffy negative phenotype [6,7]. For decades, it was believed that Duffy negative individuals were completely refractory to *P. vivax* infection. However, since 2006, multiple reports of Duffy negative infections by *P. vivax* have been made [8–13]. In Ethiopia, previous studies have shown that the prevalence of *P. vivax* among Duffy negative individuals varied by study sites, ranging between 4-12% based on endemicity and proportion of Duffy negative individuals in the general population [14–17].

Although never demonstrated, it was suggested that *PvDBP* gene duplication was necessary to enable *P. vivax* invasion in Duffy negative reticulocytes [11,18,19]. *PvDBP* gene duplication would enable *P. vivax* to bind to either an unknown receptor of a *DARC* independent pathway with a lower affinity to *PvDBP* than *DARC*, or to small amounts of *DARC* on Duffy negative reticulocytes. However, *PvDBP* gene duplication has also been shown to be involved in the response of *P. vivax* to host anti-*PvDBP* immunity. Whether an interplay between host immunity and Duffy receptor variability leads to the selection of *PvDBP* gene duplication is currently unknown. The distribution and frequency of *PvDBP* gene duplications across different geographic areas and their relationship to Duffy genotype remain poorly characterized, including in Ethiopia. In this study, we aimed to (i) assess the prevalence and distribution of malaria species across three Ethiopian regions; (ii) characterize the distribution of Duffy genotypes in the populations; and (iii) evaluate the frequency and potential functional implications of *PvDBP* gene duplications. These findings are of key epidemiological importance and provide valuable insights into the host-parasite genetic interactions that shape malaria susceptibility in Ethiopia.

## Methods

### Participant’s enrolment, study site and sample collection

A total of 1,154 samples were collected from febrile patients seeking treatment in health facilities located in three regions of Ethiopia, including Gambela (n = 555), Oromia (n = 303), and SNNPR (n = 296), between 2020 and 2021 (Fig 1). Venous blood samples were collected in EDTA tubes from these participants. Plasma and red blood cell pellets were separated and immediately frozen at -20°C. Frozen blood products were shipped to Institut Pasteur in Cambodia (IPC) for analysis. Scientific and ethical clearance was obtained from the Ethics Review Board of the Ethiopian Public Health Institute (IRB/531/2023), Ethiopia. Written informed consent/assent for study participation was obtained from all participants of the study and parents/guardians (for minors under 18 years old).

**Fig 1.**
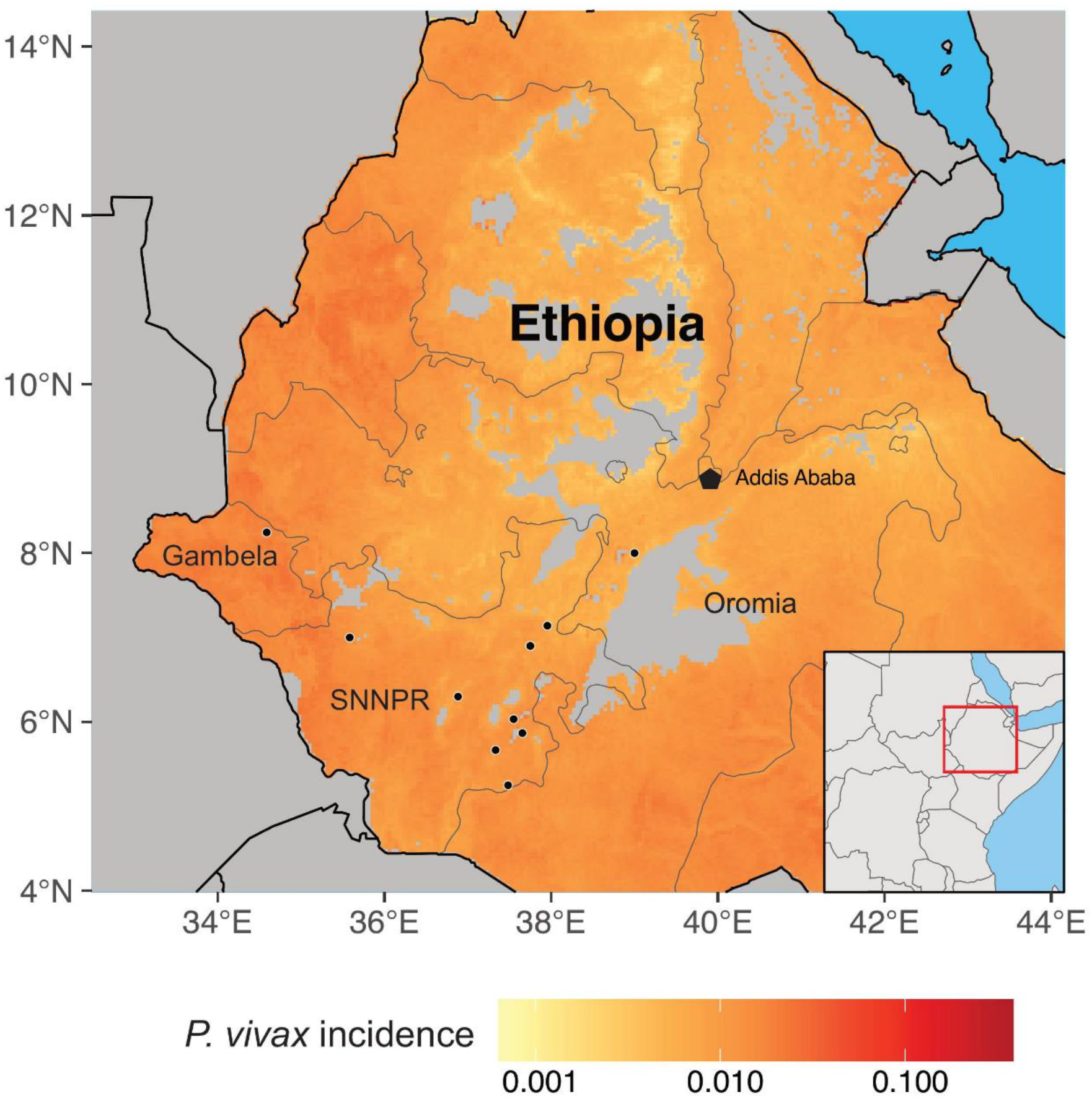
Map showing the geographical distribution of the sample sites and *P. vivax* incidence in Ethiopia.

### *Plasmodium* diagnostics and Duffy promoter genotyping

DNA was extracted from frozen whole blood samples using the DNeasy extraction kit (QIAGEN) following the manufacturer’s instructions. Detection of *Plasmodium* parasites was done on all 1,154 samples using a genus-specific real-time PCR assay targeting the *cytB* gene following established protocols [20]. *Plasmodium* species were identified by species-specific nested RT-PCR targeting the *cytB* gene in all samples positive for *Plasmodium* genus PCR following established protocols [23].

Duffy genotyping was performed using a Taqman-based multiplex qPCR assay for allelic discrimination to detect the point mutation (c.1-67T>C; rs2814778) in the GATA-1 transcription factor binding site of the *DARC* gene. The following primers (forward: 5’-GGCCTGAGGCTTGTGCAGGCAG-3’; reverse: 5’-CATACTCACCCTGTGCAGACAG-3’) and dye-labeled probes (FAM-CCTTGGCTCTTA[*C*]CTTGGAAGCACAGG-BHQ; HEX-CCTTGGCTCTTA[*T*]CTTGGAAGCACAGG-BHQ) were used. Each PCR contained 5 μl TaqMan Fast Advanced Master mix (Thermo), 1 μl DNA template, and 0.5 μl of each primer (10nM), and 0.5 μl of each probe (10nM). The reactions were performed with an initial denaturation at 95°C for 2 minutes, followed by 45 cycles of denaturation at 95°C for 3 seconds and annealing at 58°C for 30 seconds. A no-template control was used in each assay. The Duffy genotypes were determined by the allelic discrimination plot based on the fluorescent signal emitted from the allele-specific probes. For *P. vivax* positive samples, a 1100-bp fragment of the *DARC* gene was further amplified using previously published primers. Each PCR contained 20μl DreamTaq PCR Mastermix, 1μl DNA template, and 0.5μl each primer. PCR conditions were 94°C for 2-min, followed by 35 cycles of 94°C for 20s, 58°C for 30s, and 68°C for 60s, followed by a 4-min extension. PCR products were purified and sequenced using the Sanger method. Chromatograms were visually inspected to determine and confirm the Duffy genotypes based on the TaqMan assays [9,17].

### *PvDBP* gene duplication determination

PvDBP gene duplication was determined using semi-nested PCR targeting the boundary of duplicated copies as previously described [21,22]. Briefly, after a primary PCR (described in ref 21), 2 μL of amplicon were used as a template following the conditions described in ref 22, using AF2+AR2 (expected size 657bp), BF+BR (643bp), and BF+AR2 (736bp or 1584bp depending on the duplication type, see ref 22). Samples with amplification for both controls (AR2+AF2 and BF+BR) were valid for analysis. If parasites carried the *PvDBP* duplication, a band of 736bp or 1584bp was observed following PCR using AR2+BF. In absence of amplification using AR2+BF, the parasite contains a single copy of the gene.

### Statistical Analysis

All statistical analyses were performed using GraphPad Prism (version 10.0.2) and R (version 4.5.0). Descriptive statistics were used to summarize demographic and clinical variables. Associations between categorical variables were assessed using Chi-square tests or Fisher’s exact tests, as appropriate. A significance level of p<0.05 was used throughout.

The Cochran-Mantel-Haenszel (CMH) Chi-square test was used to evaluate associations between three categorical variables while controlling for stratification. When significant associations were identified, post-hoc comparisons were performed using Chi-square or Fisher’s exact tests, with Bonferroni correction applied to adjust for multiple testing. Chi-square or Fisher’s exact tests were also used to screen independent variables for inclusion in multivariable models. Three different outcomes were modeled:

(1) *Plasmodium* PCR positivity, and (2) *Plasmodium* species detected (binary and multinomial outcomes). The first two outcomes were analyzed using multivariable logistic regression, while the third outcome (with multiple species categories) was analyzed using multinomial logistic regression. For each model, backward stepwise selection was used to identify the most parsimonious set of predictors. Variables were sequentially removed from the full model, and model fit was evaluated at each step. Final models were selected based on statistical significance and model performance metrics.

## Results

### Molecular detection of *Plasmodium* species by study sites

The overall *Plasmodium* prevalence was 46% (95% CI 44-49) with significant differences between the three regions: 68% (95% CI 63-73), 43% (95% CI 37-49), and 36% (95% CI 32-40) in Oromia, SNNPR, and Gambela, respectively (p<0.0001) (Table 1, Fig 2). Among the four *Plasmodium* species detected, *P. falciparum* was the most frequent with 49% (261/535) of infections, followed by *P. falciparum/P. vivax* co-infections (26%, 140/535) and *P. vivax* mono-infections (23%, 123/535). Co-infections of *P. falciparum* or *P. vivax* with *P. ovale* and *P. malariae* were detected in 1.3% (7/535) of infected samples. A triple *P. falciparum, P. vivax* and *P. malariae* co-infection was detected in two samples (0.4%, 2/535), and single *P. ovale* and *P. malariae* infections were detected in one sample for each species (0.2%, 1/535). The distribution of *P. falciparum*, mixed *P. falciparum*/*P. vivax* and *P. vivax* infections varied among the three regions (p<0.0001). Similar proportions of *P. falciparum* infections were observed in SNNPR (55%, 70/127) and Oromia (57%, 118/206) while Gambela had a lower proportion of this species (36%, 73/202). Conversely, mixed *P. falciparum*/*P. vivax* infections were most common in Gambela (37%, 75/202) compared to Oromia (22%, 45/206) and SNNPR (16%, 20/127). The proportion of *P. vivax* infections was not significantly different between the three regions with 28% (36/127), 22% (44/202) and 21% (43/206) in SNNPR, Gambela and Oromia, respectively (p=0.6291) (Table 1; Fig 2).

**Table 1.** *Plasmodium* prevalence and proportions of species identified in the different study sites.

|  | Overall<br>n=1154 | Gambela<br>n=555 | Oromia<br>n=303 | SNNPR<br>n=296 | P value* |
| --- | --- | --- | --- | --- | --- |
| Plasmodium prevalence % (95% CI, n) | 46% (44-49, 535) | 36% (33-40, 202) | 68% (63-73, 206) | 43% (37-49, 127) | <0.0001 |
| Plasmodium species proportions (% , n) |  |  |  |  |  |
| Pf | 49% (261) | 36% (73) | 57% (118) | 55% (70) | < 0.0001 |
| Pf/Pv | 26% (140) | 37% (75) | 22% (45) | 16% (20) | < 0.0001 |
| Pv | 23% (123) | 22% (44) | 21% (43) | 28% (36) | 0.6291 |
| Pf/Po | 0.6% (3) | 1.5% (3) | - | - | - |
| Pv/Pm | 0.4% (2) | 0.5% (1) | - | 0.8% (1) | - |
| Pf/Pv/Pm | 0.4% (2) | 1% (2) | - | - | - |
| Pf/Pm | 0.2% (1) | 0.5% (1) | - | - | - |
| Pv/Po | 0.2% (1) | 0.5% (1) | - | - | - |
| Po | 0.2% (1) | 0.5% (1) | - | - | - |
| Pm | 0.2% (1) | 0.5% (1) | - | - | - |

\* Chi-square comparison of the three sites. *Pf*: *P. falciparum*, *Pv*: *P. vivax*, *Pm*: *P. malariae*, *Po*: *P. ovale*

**Fig 2.**
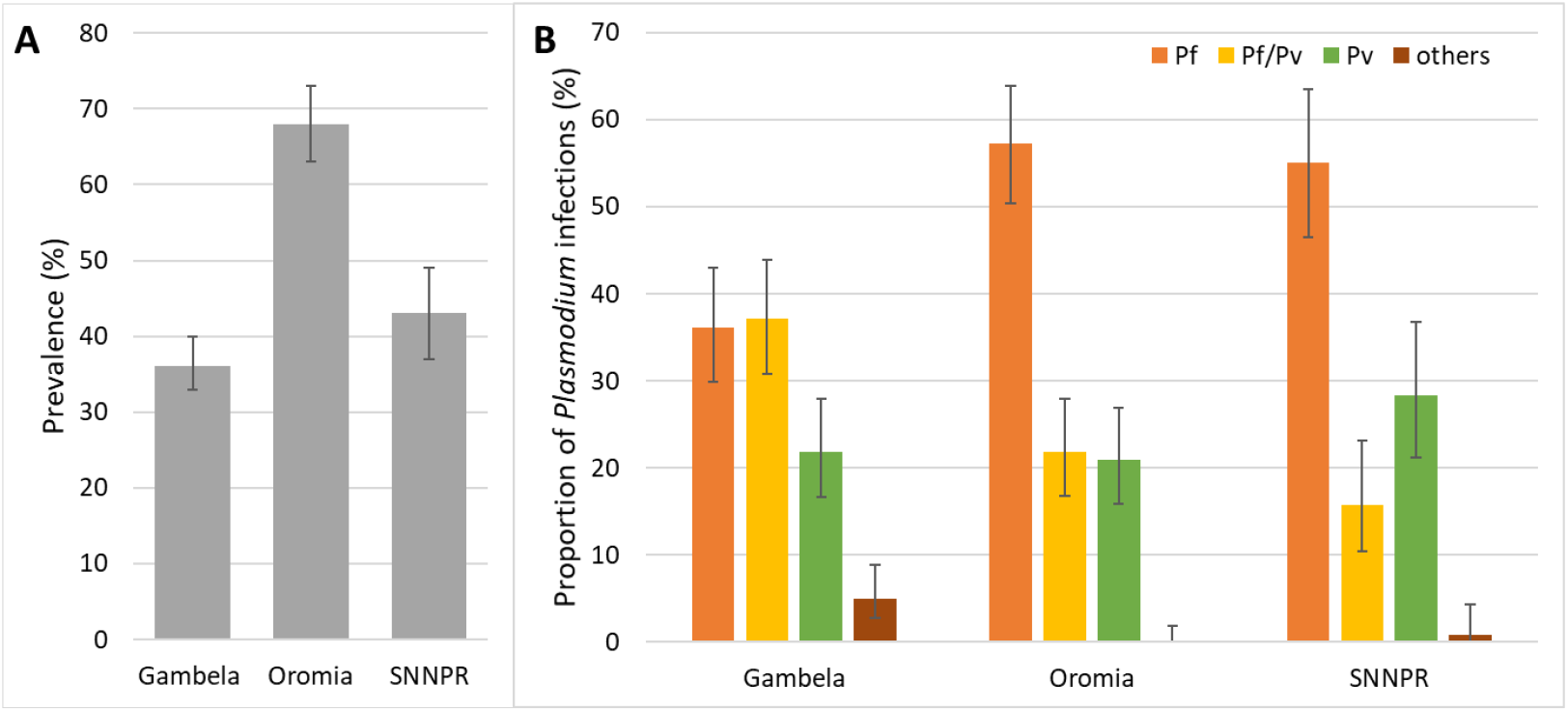
Prevalence of *Plasmodium* infections across the three regions. (A). Proportion of *Plasmodium* species in each region (B). Pf: *P. falciparum*, Pv: *P. vivax. Pf/Pv*: *P. falciparum* and *P. vivax* co-infection. Error bars represent 95% CI.

### Demographic differences in malaria prevalence

Complete demographic data were collected for 887 out of 1,154 participants. The overall sex distribution was 54% male (477/887) and 46% female (410/887). Sex distribution differed significantly across regions (p<0.0001), with a higher proportion of males in Oromia (73%) compared to more balanced distributions in Gambela (48% male) and SNNPR (50% male) (Table 2). Participants aged 20-29 years comprised the largest age group (46%, 95% CI 42-49), with age distributions also varied significantly by region (p<0.0001). Gambela had the highest proportion of individuals aged 20-29 (54%, 95% CI 49-58), followed by Oromia (41%, 95% CI 35-48) and SNNPR (21%, 95% CI 15-28).

**Table 2.** Demographic characteristics of patients enrolled in the study.

|  | **Total** | **Gambela** | **Oromia** | **SNNPR** | **P value\*** |
| --- | --- | --- | --- | --- | --- |
| Patients enrolled n | 1154 | 555 | 303 | 296 |  |
| With demographic data n | 887 | 555 | 191 | 141 |  |
| **Sex % (95% CI, n)** |  |  |  |  |  |
| Male | 54% (50-57, 477) | 48% (44-52, 267) | 73% (66-79, 139) | 50% (42-58, 71) | <0.0001 |
| Female | 46% (43-50, 410) | 52% (48-56, 288) | 27% (21-34, 52) | 50% (42-58, 70) |  |
| **Age group (years) n (% , 95% CI)** |  |  |  |  |  |
| <10 | 5% (4-7, 47) | 4% (3-6, 23) | 4% (2-8, 8) | 11% (7-18, 16) | <0.0001 |
| 10-19 | 18% (16-21, 160) | 13% (10-16, 72) | 28% (22-35, 54) | 24% (18-32, 34) |  |
| 20-29 | 46% (42-49, 405) | 54% (49-58, 297) | 41% (35-48, 79) | 21% (15-28, 29) |  |
| >29 | 31% (28-34, 275) | 29% (26-34, 163) | 26% (20-33, 50) | 44% (36-52, 62) |  |

\* Chi-square comparison of the three sites

Conversely, individuals older than 29 were more common in SNNPR (44%, 95% CI 36-52) than in Gambela (29%, 95% CI 26-33) or Oromia (26%, 95% CI 20-33).

Among the 887 study participants with demographic data, the prevalence of *Plasmodium* infection was 47% (95% CI 44-50), similar to the prevalence observed in the full cohort (46%, 44-49, p=0.8087) (Table 3). Prevalence was significantly higher in males (60%, 95% CI 55-64) compared to females (32%, 95% CI 28-37, p<0.0001). Malaria prevalence remained significantly different by regions among these participants (p<0.0001). Individuals over 29 years had the lowest prevalence (32%, 95% CI 27-38) while similar prevalence were observed in younger groups and ranged from 49% (95% CI 35-63) in patients aged less than 10 years old to 54% in both patients aged 10 to 19 years old (95% CI 46-61) and patients aged 20 to 29 years old (95% CI 49-59, p<0.0001) (Table 3).

**Table 3.** Demographic characteristics associated with *Plasmodium* infections.

|  |  | Number of samples | Plasmodium prevalence<br>% (95% CI, n) | P value |
| --- | --- | --- | --- | --- |
| **Overall** | All patients enrolled | 1154 | 46% (44-49, 535) | 0.8087 |
|  | With demographic data | 887 | 47% (44-50, 416) |  |
| **Regions** | Gambela | 555 | 37% (33-40, 202) | <0.0001 |
|  | Oromia | 191 | 84% (78-89, 161) |  |
|  | SNNPR | 141 | 38% (30-46, 53) |  |
| **Sex** | Male | 477 | 60% (55-64, 284) | <0.0001 |
|  | Female | 410 | 32% (28-37, 132) |  |
| **Age group (yrs)** | <10 | 47 | 49% (35-63, 23) | <0.0001 |
|  | 10-19 | 160 | 54% (46-61, 86) |  |
|  | 20-29 | 405 | 54% (49-59, 219) |  |
|  | >29 | 275 | 32% (27-38, 88) |  |

A Cochran-Mantel-Haenszel test revealed a significant interaction between sex and age group on infection prevalence in Gambela (p<0.0001). Post-hoc analysis with Bonferroni correction showed that among individuals aged 20-29, males had a significantly higher prevalence (63%, 95% CI 55-70) than females (27%, 95% CI 20%-35%, p<0.0001) (S1 Fig). In contrast, no significant association was found between sex and malaria prevalence in SNNPR and Oromia after controlling for age.

### Distribution of Duffy genotypes and malaria infections

Duffy genotypes were obtained from 721 samples, including most of the *Plasmodium* positive samples (96%, 516/535) and a random selection of 205 *Plasmodium* negative samples. Overall, similar prevalence of Duffy negative homozygous (referred to as Duffy negative, 41%, 95% CI 37-44) and Duffy positive heterozygous (referred to as heterozygous, 44%, 95% CI 40-47) individuals was observed, while prevalence of homozygous Duffy positive individuals (referred to as Duffy positive,16%, 95% CI 13-19) was lower (p<0.0001) (Table 4). The Duffy genotype distribution varied significantly across the three regions (p=0.0018), with the highest prevalence of Duffy negative individuals in Gambela (48%, 95% CI 42-53) and the lowest in SNNPR (30%, 95% CI 24-37). In contrast, the highest prevalence of heterozygous (50%, 95% CI 43-57) and Duffy positive individuals (20%, 95% CI 15-26) was observed in SNNPR (Table 4; Fig 3).

**Table 4.**
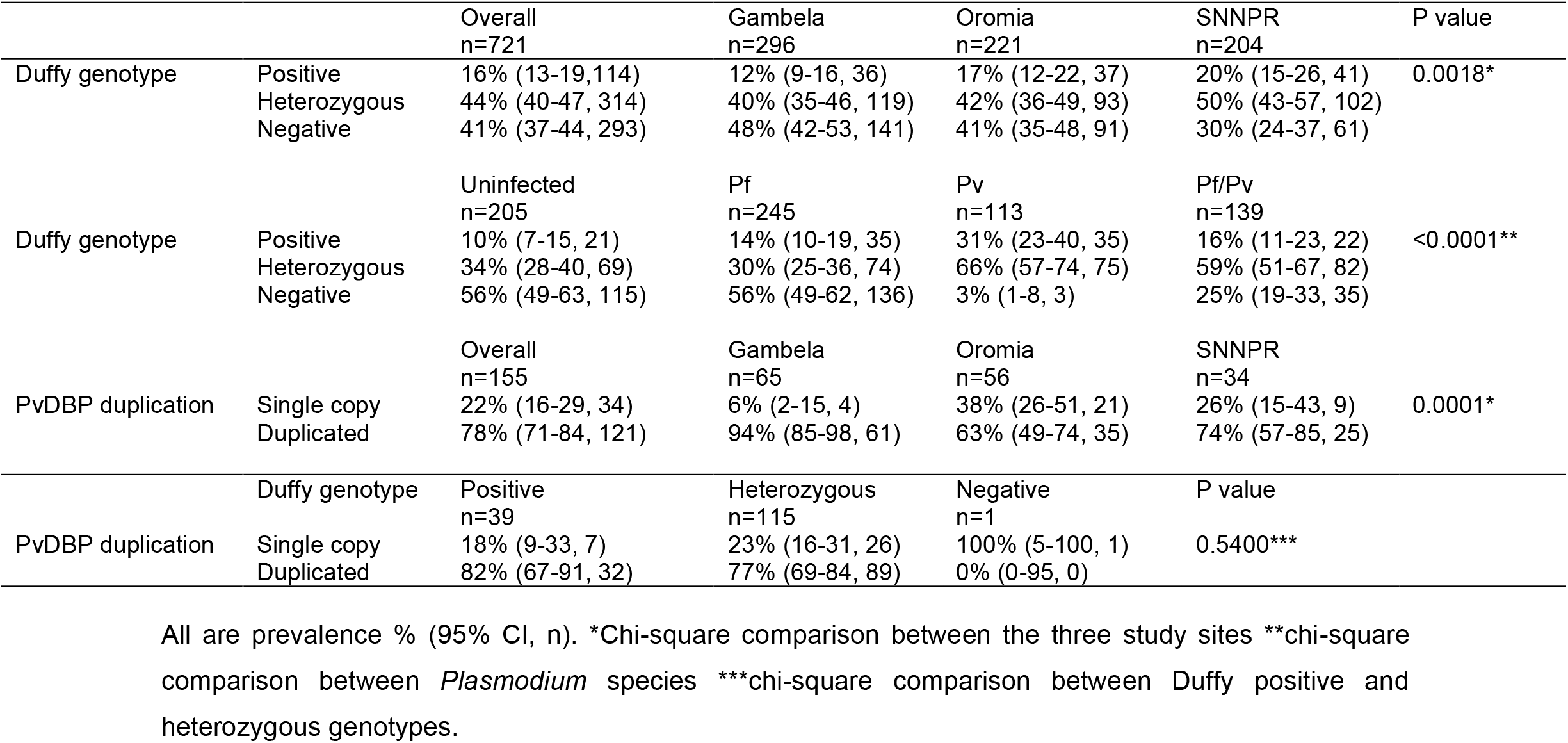
Prevalence of Duffy genotypes and PvDBP gene duplication.

**Fig 3:**
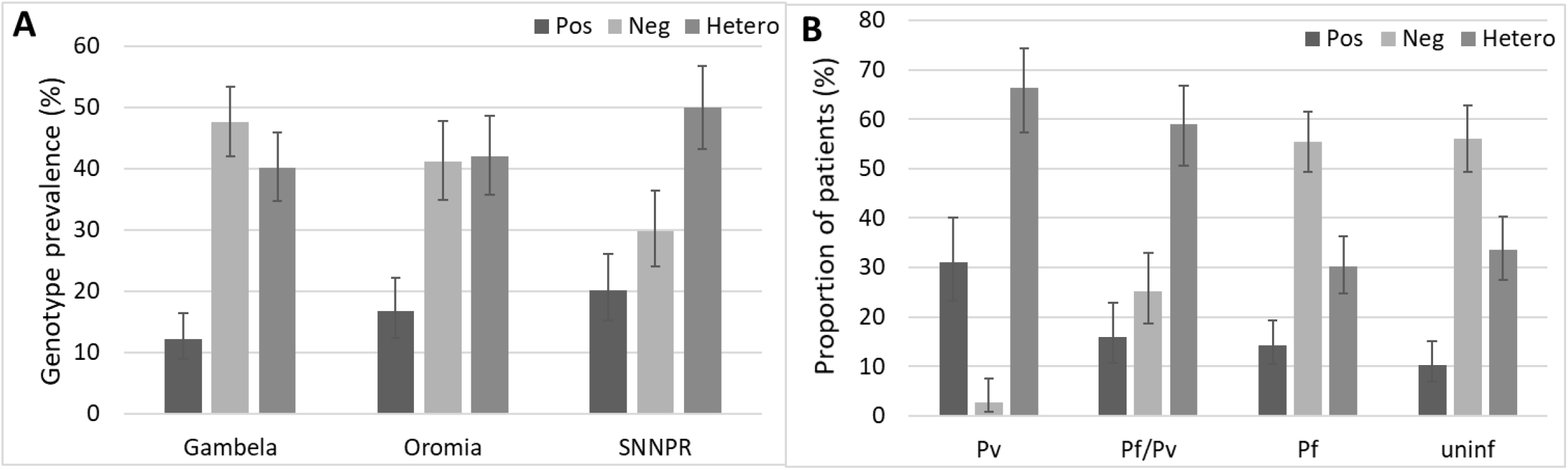
Prevalence of Duffy genotypes in the three study sites. (A). Proportion of patients Duffy positive (Pos), negative (Neg), and heterozygous (hetero) among those infected by *Plasmodium vivax* (Pv), mixed *P. falciparum/P. vivax* (*Pf/Pv*), *P. falciparum* (*Pf*), and uninfected (uninf) (B). Error bars represent 95% CI.

There was a significant association between Duffy genotypes and *Plasmodium* species (p<0.0001). No significant difference was observed in Duffy genotype distribution between uninfected patients and those infected with *P. falciparum* (p=0.3884). As expected, *P. vivax* mono-infections were rare among Duffy negative individuals (3%, 95% CI 1-8), compared to uninfected individuals (56%, 95% CI 49-63, p*<*0.0001). Interestingly, among individuals with mixed *P. falciparum*/*P. vivax* infections, 25% (95% CI 19-33) were Duffy negative, a significantly higher proportion than among *P. vivax* cases (p<0.0001), although lower than among uninfected individuals (p<0.0001) and *P. falciparum* infected ones (56%, 95% CI 49-62, p<0.0001) (Fig 3, Table 4).

### PvDBP gene duplication analysis

A subset of 263 *P. vivax* infected samples, either as mono-infections or in mixed infections, were tested for PvDBP gene duplication. Valid results (with both PCR controls leading to the expected amplicons) were obtained for 155 samples. Overall, PvDBP gene duplication was detected in 78% (95% CI 71-84) of these samples, and its prevalence was different between regions (p=0.0001) (Table 4). The proportion of duplication was significantly higher in Gambela (94%, 95% CI 85-98) compared to SNNPR (74%, 95% CI 57-85, p=0.0045) and Oromia (63%, 95% CI 49-74, p<0.0001). No significant difference was observed between Oromia and SNNPR. In addition, the proportion of gene duplication was similar between parasites collected from Duffy positive homozygous (82%, 95% CI 67-91) and heterozygous individuals (77%, 95% CI 69-84, p=0.6543). All samples carried the Cambodian-type PvDBP duplication, with none showing the Malagasy-type. A single *P. vivax* infection from a Duffy negative individual showed no duplication.

## Discussion

This study provides a molecular and epidemiological characterization of *Plasmodium* infections in febrile, treatment-seeking patients from three distinct regions of Ethiopia. The overall malaria prevalence of 46% underscores the continued public health burden of malaria in the region, with substantial variation across geographic areas. Notably, Oromia exhibited the highest prevalence, largely driven by its extensive lowland ecology covering approximately 85% of the area, which facilitates malaria vectors’ reproduction and parasite development [23–26]. The effectiveness of vector control in Oromia is undermined by the inconsistent use of insecticide-treated nets (ITNs) and long-lasting insecticidal nets (LLINs), which is further compounded by the emergence of insecticide resistance among mosquito populations [27–29]. In contrast, in Gambela, despite its hot and humid climate with seasonal rainfall patterns favoring vectors breeding [30], exhibited lower malaria prevalence compared to Oromia. This can be explained by differences in health-seeking behavior, the implementation of malaria prevention strategies, and malaria diagnostic capacity [31]. Moreover, limited healthcare access and potential underreporting of malaria cases remain a challenge due to the presence of mobile pastoralist communities who are difficult to reach by standard health services [32]. The lowest malaria prevalence recorded in SNNPR can be attributed to its predominantly highland geography, where cooler temperatures limit malaria transmission [33]. Additionally, SNNPR benefits from a strong health extension worker program and relatively high ITN coverage in high-risk areas, sustaining prevention efforts [28]. These findings highlight the heterogeneous transmission dynamics in Ethiopia, influenced by the interaction of ecological factors, healthcare access, and vector control effectiveness across the three regions.

We observed a significantly higher malaria prevalence among male young adults than other demographic groups in Gambela. This could be the result of occupational exposure and behavioral factors. The lack of age/sex association with malaria prevalence in SNNPR and Oromia could suggest context-dependent drivers that merit further exploration or an insufficient sample size to identify such an association. Notably, participants recruited for this study may also be skewed by sex and age. Males, particularly young adults, were overrepresented in the enrolled cohort, especially in Gambela. While this may partly be due to population-level disease risk, it could also indicate behavioral and sociocultural factors influencing health-seeking behavior. Underrepresentation of women, particularly of reproductive age, may underestimate the burden of malaria in this group and obscure sex-specific disease dynamics [34]. This raises the possibility that surveillance data relying on health facility-based sampling may underrepresent malaria burden among women, particularly if they face barriers to accessing healthcare. More inclusive recruitment strategies are warranted to ensure balanced representation across demographic strata and to avoid skewed interpretations of transmission dynamics.

Among the *Plasmodium* species identified, *P. falciparum* was predominant. However, a significant proportion of *P. vivax* infections and more notably *P. falciparum*/*P. vivax* co-infections were also detected. Less common *Plasmodium* species, including *P. malariae* and *P. ovale* were also detected in the regions, either as single or co-infections. These infections are often overlooked due to diagnostic challenges and the tendency for microscopy to miss low-density infections [35]. This reinforces the need for multi-species diagnostic tools and treatment regimens, particularly given the different biological characteristics and relapse potential of *P. vivax*.

Our findings confirm a strong association between Duffy blood group polymorphisms and the risk of *P. vivax* infection. As expected, Duffy-negative individuals were less susceptible to *P. vivax* infections. Interestingly, we show here that a significant proportion of Duffy-negative individuals were co-infected with *P. falciparum* and *P. vivax*. While the exact mechanism remains unclear, it is plausible that *P. falciparum* induced hemolysis and subsequent reticulocyte production create a favorable niche for *P. vivax*, which preferentially invades young red blood cells. This observation warrants further investigation into potential facilitation by *P. falciparum* and could have implications for the evolutionary forces that drove the selection of Duffy-negative genotypes as previously suggested [36].

Our findings indicated a high prevalence of PvDBP gene duplication in *P. vivax* isolates, particularly in Gambela where the prevalence of Duffy-negative individuals was the highest and where mixed *P. falciparum*/*P. vivax* infections were the most prevalent. Although duplication was not observed in the single Duffy-negative individual, the high frequency of duplication in this region suggests that variability in Duffy receptor may exert significant pressure to select for parasite genomic adaptation, in addition to other evolutionary drivers such as host immune pressure. Additional studies are needed to evaluate the role of this gene duplication in the invasion of Duffy-negative reticulocytes by *P. vivax*. Our data indicate that all parasites with the duplication carried the Cambodian type, and no Malagasy type duplication was detected. This finding is consistent with previous studies conducted in Ethiopia, where most *P. vivax* infections were associated with the Cambodian-type duplication [19,22,37,38]. Further population genetic studies are needed to better understand the mechanisms underlying these patterns. Moreover, future research should prioritize the collection of more comprehensive epidemiological, clinical, and genetic data from diverse geographic regions. These initiatives will facilitate a better understanding of the impact of PvDBP gene duplication on parasite fitness, disease severity, and the ability to infect Duffy-negative individuals.

## Data Availability

All data produced in the present study are available upon reasonable request to the authors.

## Author contributions

SM, EL, and JP conceived and designed the study; TTS, AA, and MT collected the samples; BT, LBF-D, BNY, VE, CT, NK, and JP collected and analyzed the data; BT, LBF-D, BNY, EL, and JP wrote and edited the paper. All authors have read and approved the final manuscript.

## Acknowledgment

We are greatly indebted to technicians and staffs at the Ethiopian Public Health Institution and from local health facilities for assisting with sample collection and blood processing; and health offices for their support and willingness to participate in this research. We would also like to express our sincere gratitude to all the patients who participated in this study.

